# Remote and unsupervised digital memory assessments can reliably detect cognitive impairment in Alzheimer’s disease

**DOI:** 10.1101/2024.01.08.24300982

**Authors:** David Berron, Emil Olsson, Felix Andersson, Shorena Janelidze, Pontus Tideman, Emrah Düzel, Sebastian Palmqvist, Erik Stomrud, Oskar Hansson

## Abstract

**INTRODUCTION:** Remote unsupervised cognitive assessments have the potential to complement and facilitate cognitive assessment in clinical and research settings.

**METHODS:** Here we evaluate the usability, validity and reliability of unsupervised remote memory assessments via mobile devices in individuals without dementia from the Swedish BioFINDER-2 study and explore their prognostic utility regarding future cognitive decline.

**RESULTS:** Usability was rated positively; remote memory assessments showed good construct validity with traditional neuropsychological assessments and were significantly associated with tau-PET and downstream MRI measures. Memory performance at baseline was associated with future cognitive decline and prediction of future cognitive decline was further improved by combining remote digital memory assessments with plasma p-tau217. Finally, retest reliability was moderate for a single assessment and good for an aggregate of two sessions.

**DISCUSSION:** Our results demonstrate that unsupervised digital memory assessments might be used for diagnosis and prognosis in Alzheimer’s disease, potentially in combination with plasma biomarkers.

## Introduction

While there has been significant progress in fluid and neuroimaging biomarkers to detect pathological changes in Alzheimer’s disease (AD), cognitive measures in health care and clinical trials were initially designed to detect overt cognitive impairment and novel developments still lag behind (Hansson, 2021; Hansson et al., 2023). This is in stark contrast to recent discoveries on the functional architecture of episodic memory and its relationship to the spatial progression patterns of AD pathology (Berron et al., 2021; Maass et al., 2019). Recent work on the functional neuroanatomy of episodic memory have highlighted memory networks in the medial temporal lobe (MTL) and the neocortex that are involved in specific memory functions and are affected in different stages of AD (Berron et al., 2021; Grothe et al., 2017; Ranganath and Ritchey, 2012; Ritchey and Cooper, 2020). Episodic memory requires pattern separation and completion processes that are primarily mediated by MTL subregions. While the dentate gyrus is involved in pattern separation (Bakker et al., 2008; Berron et al., 2016) and reduces memory interference between similar events, the hippocampal Cornu Ammonis 3 (CA3) mediates pattern completion processes in interplay with neocortical regions (Grande et al., 2019). Within the hippocampal entorhinal circuitry there exist partly segregated pathways where object information is primarily provided from the perirhinal cortex via the anterior-lateral entorhinal cortex. Spatial information via the parahippocampal cortex is transferred additionally through the posterior parts of the entorhinal cortex (Berron et al., 2019, 2018; Grande et al., 2022; Maass et al., 2019, 2015; Schröder et al., 2015). Taken together, there is converging evidence that short-term mnemonic discrimination of object and scene representations, in addition to long-term memory, is impaired in the predementia stages of AD (Grande et al., 2021).

In addition, traditional neuropsychological assessment suffers from significant limitations such as high participant burden and impracticality of implementing test formats such as frequent test repetitions or long-term delay formats. Thus, traditional neuropsychological assessments become increasingly difficult for clinical trials in preclinical and early-stage symptomatic AD populations which gradually implement decentralized clinical trial structures for case-finding and monitoring, for example TRAILBLAZER-ALZ 3 (ClinicalTrials.gov Identifier: NCT05026866). In this context, unsupervised and remote digital cognitive assessments via smartphones and tablets offer a promising avenue to improve case-finding, monitoring and prognosis in both clinical trial and health-care settings (Nicosia et al., 2022b; Öhman et al., 2021, 2022; Papp et al., 2021). The aim of the present study was to evaluate the feasibility and usability of unsupervised and remote digital memory assessments, their construct validity in reference to traditional neuropsychological assessments, their retest reliability as well as their relationship with fluid and imaging biomarkers of AD. To that end, we implemented two non-verbal memory tasks based on recent insights into the functional anatomy of episodic memory, available on the neotiv digital platform (https://www.neotiv.com/en) (Berron et al., 2022; Öhman et al., 2022), in a subset of the Swedish BioFINDER-2 study.

## Methods

### Recruitment into smartphone-based add-on study

A total of 187 individuals who participated in the 7 Tesla MRI study arm of the Swedish BioFINDER2 study were offered participation in biweekly smartphone-based remote cognitive assessments across a 12-month period. 130 individuals (70%) gave written informed consent to participate.

A brief pen-and-paper questionnaire about experience with smartphones and mobile apps was completed and most participants downloaded and installed the neotiv app directly after the MRI session and completed the first phase of the first digital memory assessment session on site. A comprehensive user manual was distributed which included download and installation instructions and those individuals that did not download and install the app on-site successfully completed it without supervision from their home. The study was approved by the Regional Ethics Committee in Lund and the Swedish Ethical Review Authority.

Following completion of the initial app-hosted cognitive assessment, participants were notified to complete memory assessments every two weeks for 12 months. Each of the assessments consisted of a two-phase session. The two phases were either two halves of mnemonic discrimination, or encoding and retrieval phases of object-in-room recall (see details of the tasks below). Each phase took less than 10 minutes. Thus, overall participants could complete up to 12 sessions for each memory task within a period of approximately 12 months. In order to minimize potential practice effects from repeated testing, 12 independent difficulty-matched parallel test versions for each memory task were used. After each task completion, participants indicated their subjective task performance and their concentration level on a 5-point scale (1=very bad, 2=bad, 3=moderate, 4=good and 5=very good), and whether they had been distracted during the task (yes/no). Of note, only the first valid session of each participant was used in these cross-sectional analyses.

Push notifications were used to notify individuals about available tasks and to remind them daily for five consecutive days in case the tasks had not been initiated. At the beginning of each test session, participants were asked to go to a quiet environment, wear their glasses if needed and to adjust their screen’s brightness to see the pictures clearly. They also received a short practice session for the initial test session as well as for all future sessions.

At the end of the study, 40 participants partook in telephone interviews to provide feedback on usability and user experience throughout the study. The full study timeline can be seen in Figure 1.

**Figure 1:**
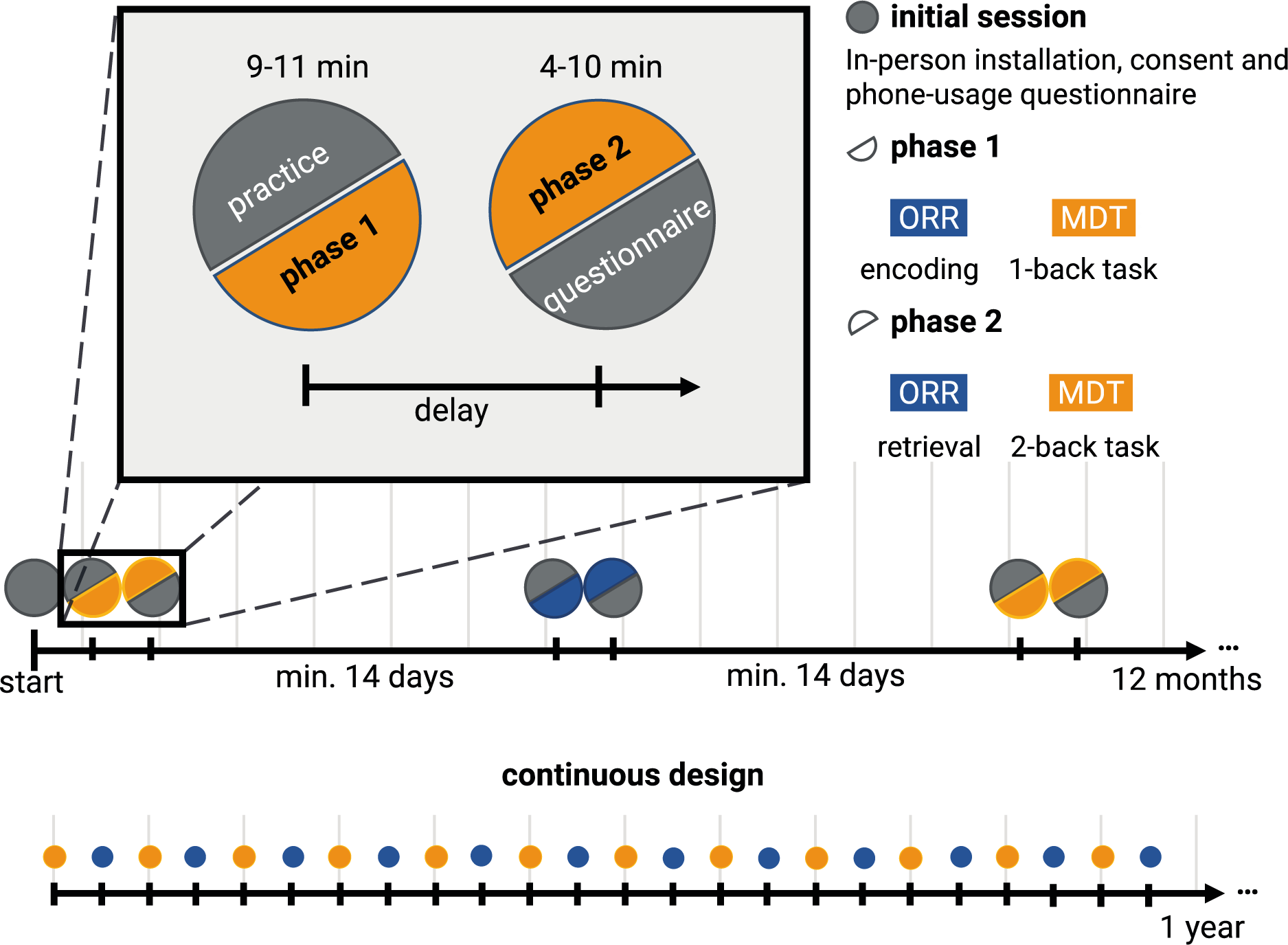
Timeline of the study protocol. Participants enlisted for a 12-month study of biweekly remote and unsupervised memory assessments of the Mnemonic Discrimination Task for Objects and Scenes (MDT-OS) and Objects-in-Room-Recall Task (ORR). In the initial in-clinic session, they gave consent, got a brief introduction, answered a short questionnaire on their phone usage, and completed the first task. Every second week, they received a short training session, followed by phase 1 of their respective task: encoding for ORR, and 1-back task for MDT-OS. After finishing phase 1, they were notified when the next phase was available, and could perform it straightaway or postpone if inconvenient; that is, there was a minimum delay of 24 h for the MDT-OS and a minimum delay of 60 minutes for the ORR, but it was often extended by the participants. Phase 2 consisted of retrieval for ORR, and 2-back task for MDT-OS. It was followed by ratings regarding concentration, distraction throughout and subjective difficulty of the task.

### Smartphone-based memory assessments

#### Mnemonic Discrimination of Objects and Scenes (MDT-OS)

In this continuous recognition task, individuals were presented with computer-generated images of various indoor objects and empty rooms, which were either exactly repeated, or slightly altered (see FigureA). Participants had to indicate whether an image was an identical repetition of a previous image (tap on a button), or had been modified (tap on the location of change). One session consisted of 64 image pairs (32 object pairs, 32 scene pairs), half of which were modified, and half of which were repeated. To optimize engagement while minimizing the subjective burden of participating, each session was split into two phases and automatically scheduled on two consecutive days with a 24-hour delay between phases. The first phase was presented as a one-back task while the second phase was presented as a two-back task. The MDT-OS has been designed to tax hippocampal pattern separation; a memory mechanism needed to discriminate between similar memories. Earlier studies using functional magnetic resonance imaging have shown that especially subregions in the human medial temporal lobe are involved in this task (Berron et al., 2019, 2018; Maass et al., 2019). The test provides a hit rate, a false alarm rate and a corrected hit rate for both the object and scene condition. The averaged corrected hit rate across the scene and the object condition is used as the main outcome measure in the following analyses.

#### Object-In-Room Recall (ORR)

In this task, participants had to memorize a spatial arrangement of two objects within a room. Following the encoding phase, a blue circle highlighted the previous position of one of the objects in the same but now empty room and the participant had to identify the correct object from a selection of three in an immediate retrieval phase (see Figure 2B). Among the three possible objects, one was the correct one that was previously shown in the now highlighted position (target), another belonged to the same room but had previously been shown at a different position (correct source distractor), and the third had previously been shown in a different room (incorrect source distractor). They learned 25 such object-scene associations. After 60 minutes, the participant was notified via push notification to complete an identical but delayed retrieval phase with a randomized stimulus order. The ORR has been designed to tax hippocampal pattern completion, a memory mechanism needed to restore full memories from partial cues (Berron et al., 2023; Grande et al., 2019). In the test, the assessment of recall is graded and allows to separate correct episodic recall from incorrect source memory. Thus, correct recall excludes the choice of an object that was present in the same room but at a different location (wrong source memory for specific location), and an object that was not present in the room but nevertheless associated with another room during encoding (wrong source memory for overall location). The test provides an immediate recall (ORR-IR; 0-25) and a delayed recall score (ORR-DR; 0-25). Here we use the ORR-DR.

**Figure 2:**
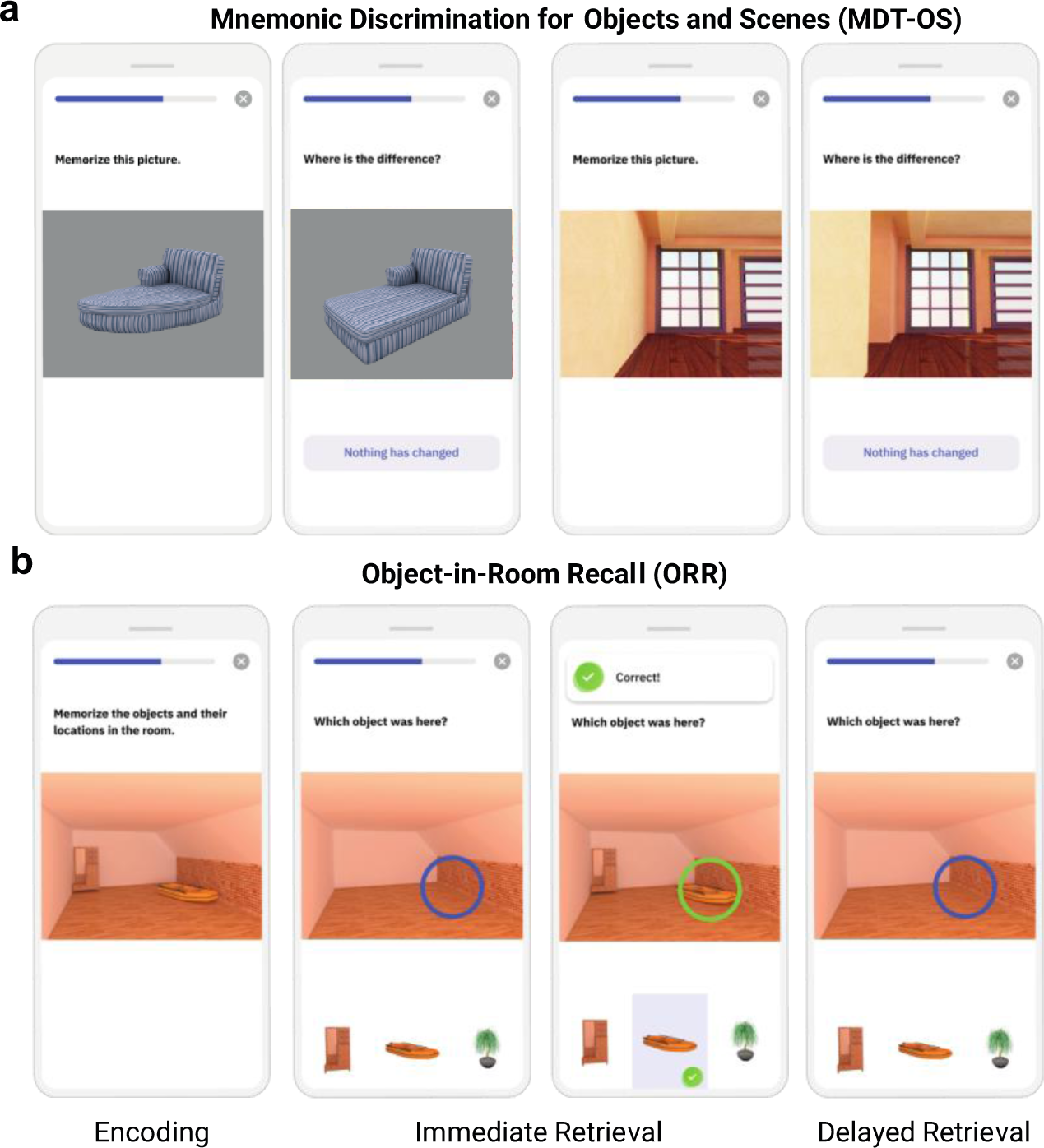
Smartphone-based memory tasks. (A) Mnemonic Discrimination Test for Objects and Scenes (MDT-OS) and (B) Objects-in-Room-Recall (ORR) test.

#### Quality Control Procedures

For this manuscript, we analyzed data up until the data release in July 2023. Recruitment of participants from the Swedish BioFINDER-2 study was performed between February 2019 and February 2022. Both memory tasks were presented in two phases, and we only included participants that had completed at least one full session, i.e., both phases. The following outcome measures were calculated from the individual trials of each memory task: For the MDT-OS, we calculated the corrected hit rate to adjust for response bias by subtracting the false alarm rate (percentage of modified images that were identified as old) from the hit rate (the percentage of repeated images that were identified as repeated). The resulting cognitive outcomes are thus the overall corrected hit rate for mnemonic discrimination (average across all object and scene trials) as well as corrected hit rates for the object and scene condition separately. For the ORR, we calculated the delayed retrieval accuracy. Regarding the sessions from the first wave, meaning each very first assessment of MDT-OS and ORR-DR, 6% of test sessions exceeded the threshold for missing responses (maximum of 25% of missing responses per session) and 17% of test sessions exceeded the maximum length of the delay period (>240 minutes) before filtering. These sessions were excluded during quality assessment. Excluded test sessions were replaced by valid subsequent sessions where possible. As a result, 67% of test sessions we report here were from the first wave and 33% from subsequent test sessions.

### CSF and plasma analysis

Cerebrospinal fluid (CSF) β-amyloid (Aβ)42 and Aβ40 concentrations were measured using the Elecsys assays (Roche Diagnostics). Participants were stratified for Aβ status using the CSF Aβ42/40 ratio with a predefined cut-off of 0.08 (Quadalti et al., 2023). Plasma levels of p-tau217 were quantified with the MSD-based immunoassay developed by Lilly Research Laboratories as previously described (Palmqvist et al., 2020).

### Imaging acquisition

#### MRI

T1-weighted images were acquired on a Siemens Prisma scanner (Siemens Medical Solutions, Erlangen, Germany) with a 64-channel head coil using an MPRAGE sequence (in-plane resolution=1×1mm^2^, slice thickness=1mm, TR=1900ms, TE=2.54ms, flip-angle=9°). Spontaneous BOLD oscillations were acquired with a gradient-echo planar sequence (eyes closed, in-plane resolution=3×3mm^2^, slice thickness=3.6mm, TR=1020ms, TE=30ms, flip-angle=63°, 462 dynamic scans, 7.85min).

### Tau and A*β*-PET

All study participants underwent positron emission tomography (PET) scans on a digital GE Discovery MI scanner (General Electric Medical Systems). Approval for PET imaging was obtained from the Swedish Medical Products Agency. For tau-PET imaging, the participants were injected with 365±20 MBq of [18F]RO948, and LIST mode emission data was acquired for each scan 70–90 min ([18F]RO948) post injection. Aβ-PET imaging was performed on the same platform 90–110 min after the injection of ∼185 MBq [^18^F]Flutemetamol (Cho et al., 2020).

### Imaging analysis

#### ROI segmentation and estimates of volume and thickness

Individual volume of the anterior and posterior hippocampus and median thickness of area 35 were defined on T1-weighted images (1x1x1mm^3^ resolution) using ASHS-T1 (Xie *et al*., 2019) and a multi-template thickness analysis pipeline (Xie et al., 2017). All subregional masks were visually assessed.

### SUVr measures

*[^18^F]RO948:* Standardized uptake value ratio (SUVr) images were calculated for area 35 using an inferior cerebellar reference region (Baker et al., 2017). Partial volume correction (PVC) was performed using the geometric transfer matrix method (Rousset et al., 1998) extended to voxel-level using region-based voxel-wise correction (Thomas et al., 2011). *[^18^F]Flutemetamol:* A cortical composite SUVr as well as a measure of early amyloid deposition in the precuneus was calculated using the whole cerebellum as reference region (Thurfjell et al., 2014).

### Traditional cognitive measures

As a measure representing episodic memory, we used the delayed 10-word list recall test from the Alzheimer’s Disease Assessment Scale—Cognitive Subscale (ADAS-Cog; Rosen et al., 1984). The learning trial of the 10 words was repeated three times. After a distraction task (Boston Naming—15 items short version), the participant was asked to freely recall the 10 words (“delayed recall”). Delayed recall was scored as number of errors (*i.e.*, 10 minus correct recalled words), so that a higher score entailed worse memory performance. For global cognition, we used the Mini-Mental State Examination (MMSE; scale from 0–30). For attentional and executive function, we used the Symbol Digit Modalities Test (SDMT, one point for every correct answer within the response time of 90 seconds) and verbal fluency (Animal fluency; number of correct animals within the response time of 60 seconds). In addition, we used the corrected hit rates for object and scene memory derived from an on-site version of the test that was completed while lying in the fMRI scanner. In the end, we calculated a composite score similar to the Preclinical Alzheimer Cognitive Composite 5 (PACC5), the modified PACC (mPACC), using the average of the z-standardized scores of ADAS delayed recall (counted twice), SDMT, MMSE and verbal fluency.

### Statistical analysis

Multiple regression analyses were carried out between PET measures, MTL subregional atrophy and unsupervised remote memory tests in R (version 4.1.2; www.r-project.org; R Core Team, 2022). All models were adjusted for age, sex and intracranial volume (for models including volumes). Results were corrected for multiple comparisons using False-Discovery Rate correction (p<0.05) where appropriate. Robust regression models were estimated using iteratively re-weighted least squares (ILRS) using the MASS package (rlm function).

We extracted participant-specific mPACC slopes from linear mixed-effects models with random intercepts and slopes (using the lme4 package for R) and mPACC score as the outcome and time (visit number) as the predictor. These participant-specific slopes were used as outcomes in a second set of linear regression models with plasma p-tau217 and remote memory measures as predictors while adjusting for age, sex and years of education. For comparison, we also fit basic models using only the covariates, without cognitive markers or biomarkers. Models were evaluated using Akaike information criterion (AIC). A difference of 2 AICs is considered significant (Burnham and Anderson, 2004). Retest reliability was assessed for two individual sessions (Session 1 vs 2; on average after 7 weeks) and for two averaged sessions (mean of Session 1 and 2 vs. mean of Session 3 and 4; on average after 13 weeks). Intraclass correlation coefficients (ICC) and their 95% confident intervals were calculated based on single rater, absolute-agreement, two-way random-effects model. We consider ICC values less than 0.5 indicative of poor reliability, values between 0.5 and 0.75 indicative for moderate reliability, values between 0.75 and 0.9 indicative for good reliability, and values greater than 0.90 indicative for excellent reliability (Koo and Li, 2016).

### Data availability

Anonymized data will be shared by request from a qualified academic investigator for the sole purpose of replicating procedures and results presented in the article and as long as data transfer is in agreement with EU legislation on the general data protection regulation and decisions by the Ethical Review Board of Sweden and Region Skåne, which should be regulated in a material transfer agreement.

## Results

### Participant sample

Here we considered the 130 study participants who opted to participate in the add-on smartphone-based study. While 20 did not complete any test session, another 11 were excluded during quality assurance as described above. As a result, 99 individuals who contributed at least one valid test session of the MDT-OS (see table 1) and 66 participants who contributed at least one valid test session of the MDT-OS and the ORR task (see supplementary table 2) could be included. Following initial filtering, we thus analyzed data from 99 participants in the MDT-OS task (48 CU Aβ–, 28 CU Aβ+, 9 MCI Aβ– and 14 MCI Aβ+) and from 66 participants in the ORR task (31 CU Aβ–, 19 CU Aβ+, 7 MCI Aβ– and 9 MCI Aβ+).

**Table 1:**
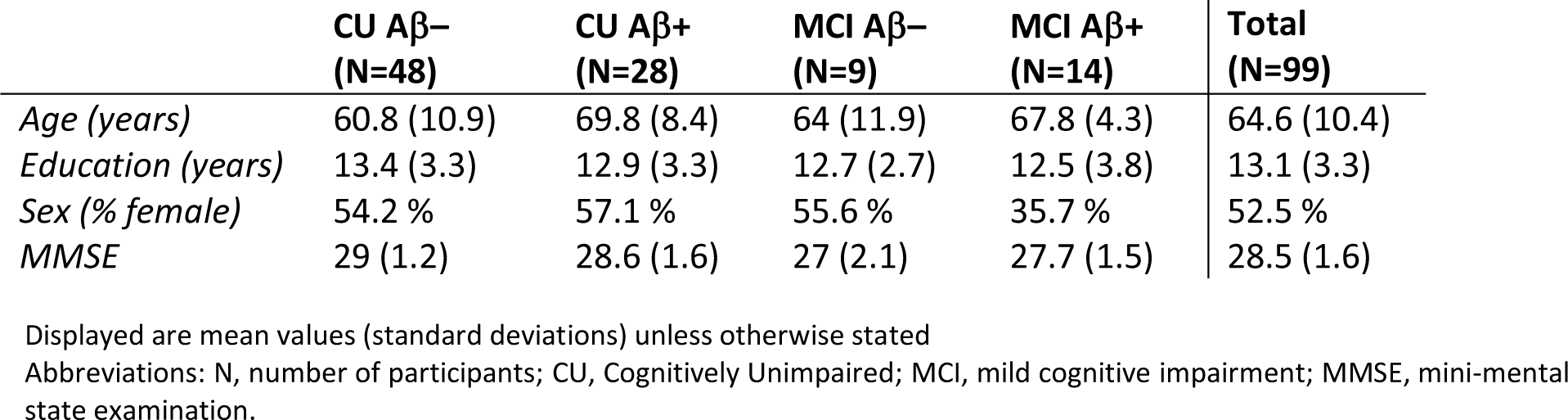
Participant demographics of the entire sample.

### Older participants show high acceptability of smartphone-based assessments

First of all, we were interested in the acceptability of the remote smartphone-based assessments. 127 participants completed a questionnaire about their phone usage. Out of those, 90% owned a smartphone, 62% said they would download apps by themselves, and 22% with assistance. Participants reported that they had been using a smartphone for 8.6 years on average (SD: 4.3 years, range: 1 – 25 years), and spend 2.3 hours per day using it (SD: 1.8 hours, range: 1–10 hours).

As reported above, 130 individuals were recruited into the add-on smartphone study (70% of those that were asked). The most frequent reason not to participate in the smartphone-based study was lack of time (too many assessments; 13%), followed by unfamiliarity with smartphone use and insecurity about their capabilities (7%), as well as not owning a smartphone with internet connection (7%). 73% did not further specify any reasons.

Across both cognitive tests, participants reported high average concentration levels (MDT-OS = 3.9; ORR = 3.9; scale 1–5, which translates to good concentration) during the task as well as moderately high subjectively rated task difficulty (MDT = 3.1; ORR = 3.4; scale 1–5, which translates to moderate subjectively rated performance). Both estimates were very similar across diagnostic groups (group average range in concentration from 3.7–4.1 and in subjective task difficulty from 2.9–3.5).

However, as expected, the second phase of the task (i.e., 2-back or delayed retrieval compared to one-back or immediate retrieval) was perceived as considerably more difficult (phase 1: 3.7; phase 2: 2.7). Across both tasks, 88% of participants reported no distractions during the task and distraction rates were higher in cognitively unimpaired individuals (on average 14% across both tasks) compared to MCI patients (on average 6% across both tasks).

Following filtering, the time between encoding and retrieval in the ORR test was on average 96 minutes (SD = 44). Mobile devices had a screen diagonal between 10.2–24.6 cm (mean 13.2 cm, SD = 3.4) indicating the use of smartphones as well as tablet computers.

Finally, we approached 40 participants for a telephone-based interview to learn about their experience with the study (see Figure 3). Over 90% of them found the tasks and instructions easy to understand, and the app easy to use. 75% said they would prefer the mobile tests over in-person paper-and-pencil tests. 80% did not experience their device’s screen as too small to see all the details in the tasks. 80% found the number of tests and their duration just right, and 82.5% rated their experience using the app 7 or higher on a 10-point scale. There were only 10% of participants who had dropped out of the study earlier but still completed the phone interview. Those rated their experience with the app lower than participants who remained in the study, and they perceived the number and duration of assessments as too long.

**Figure 3:**
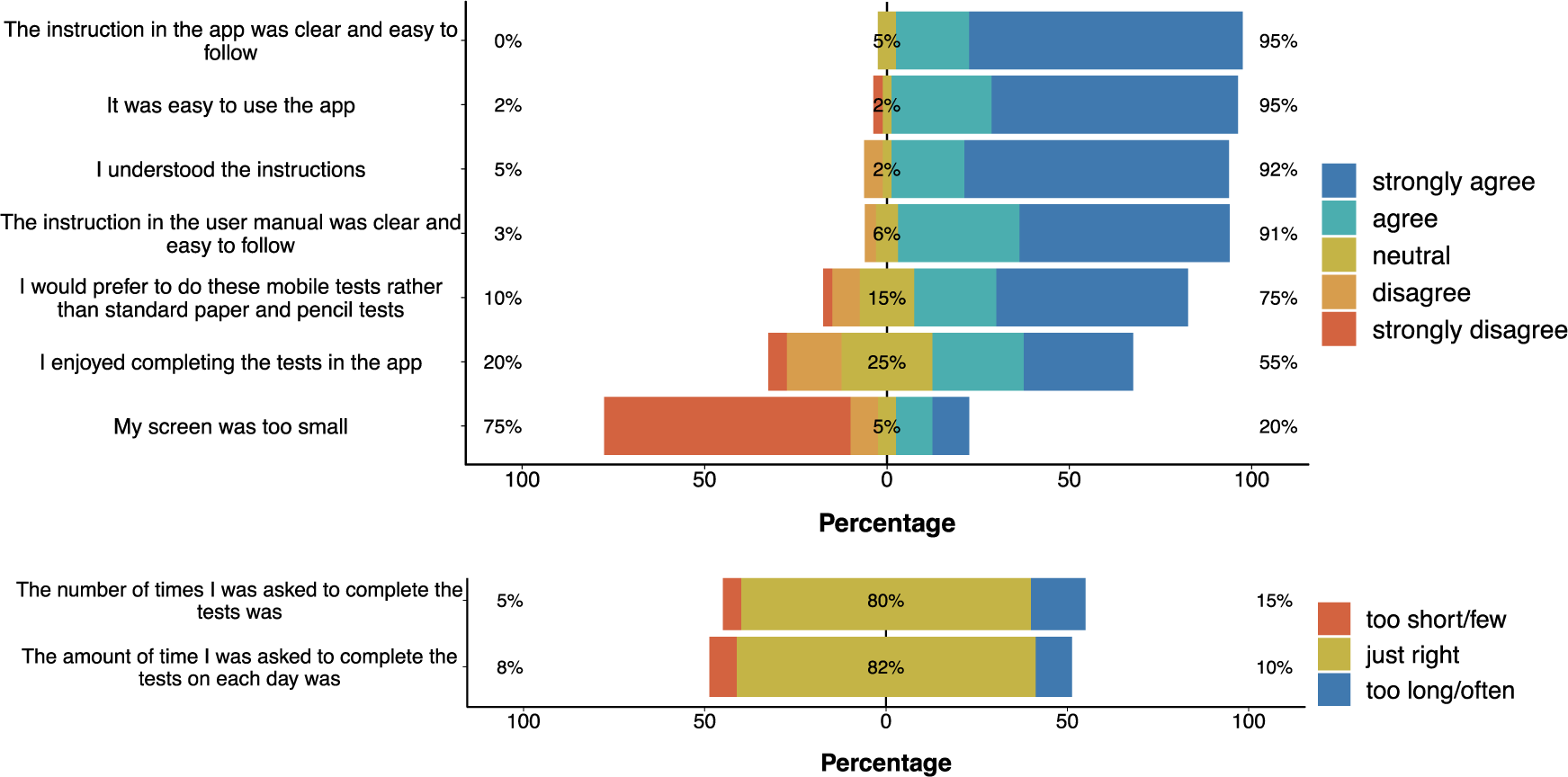
Acceptability and User experience. Results from telephone-based interviews with 40 participants focusing on their overall experience with the remote and unsupervised study.

### Unsupervised memory assessments via mobile devices reflect in-clinic cognitive scores

Next, we were interested whether the performance in the unsupervised and remote memory assessments was associated with traditional in-clinic cognitive measures. In order to understand the relationship with demographics and potential confounds, we first calculated multiple regression models with age, sex, and years of education to identify the associations with both memory paradigms. In addition, we included time-to-retrieval, i.e., the time period between memory encoding and retrieval, as a covariate for the ORR. For MDT-OS, age and sex were significant predictors for the object, and only age was a significant predictor for the scene condition where higher age and female sex were associated with worse task performance (MDT-O: β_age_ = -0.009, *p*=<.001; β_sex_ = -0.08, *p* =0.03; β_education_ = 0.005, *p* =0.34; MDT-S: β_age_ = -0.008, *p*<.001; β_sex_ = -0.06, *p*=0.13; β_education_ = 0.008, *p*=0.186). For the ORR, again only age was associated with task performance (β_age_ = -0.23, *p*<.001; β_education_ = 0.09, *p*=0.555; β_sex_ = 0.39, *p*=0.695; β_delay_ = -0.09, *p*=0.890).

Next, we assessed the construct validity of the outcomes of the unsupervised memory assessments by analyzing the relationship with several in-clinic and supervised cognitive assessments (see Figure 4). This included an in-scanner version of the MDT-OS which was completed by the same participants during an fMRI scan. While this was a similar task (mnemonic discrimination of objects and scenes; (Berron et al., 2019, 2018; Maass et al., 2019), task administration (controller button presses instead of a touch screen) and environment (lying in an MRI scanner vs. completing tasks at home) differed compared to the remote unsupervised setting. Pearson correlation coefficients revealed a strong relationship between the MDT-OS corrected hit rate across both settings (*r*=0.63, *p*<.001; MDT-O: *r*=0.43, *p*<.001; MDT-S: *r*=0.49, *p*<.001). Next, we were interested whether MDT-OS and ORR-DR were associated with an in-clinic measure of memory (ADAS delayed recall) and a composite score that has been shown to be sensitive to early AD (mPACC) as well as the Symbol Digit Modalities Test (SDMT), verbal fluency and the Mini-Mental State Examination (MMSE). Due to ceiling effects of ADAS delayed recall and the MMSE in early Alzheimer’s disease, we assessed Spearman rank correlation rho. All measures were significantly associated with the MDT-OS corrected hit rates (*ADASdelayed*: ρ=0.48, *p*<.001; MDT-O: ρ=0.53, *p*<.001; MDT-S: ρ=0.33, *p*<.001; *MMSE*: ρ=0.44, *p*<.001; MDT-O: ρ=0.35, *p*<.001; MDT-S: *r*=0.41, *p*<.001; *VerbalFluency*: *r*=0.55, *p*<.001; MDT-O: *r*=0.49, *p*<.001; MDT-S: *r*=0.49, *p*<.001; *SDMT*: *r*=0.59, *p*<.001; MDT-O: *r*=0.55, *p*<.001; MDT-S: *r*=0.50, *p*<.001). Similarly, ORR delayed retrieval performance was most strongly associated with ADAS delayed recall but also with all other outcomes (*ADASdelayed*: ρ=0.66, *p*<.001; *MMSE*: ρ=0.41, *p*<.001; *VerbalFluency*: *r*=0.54, *p*<.001; *SDMT*: *r*=0.58, *p*<.001).

**Figure 4:**
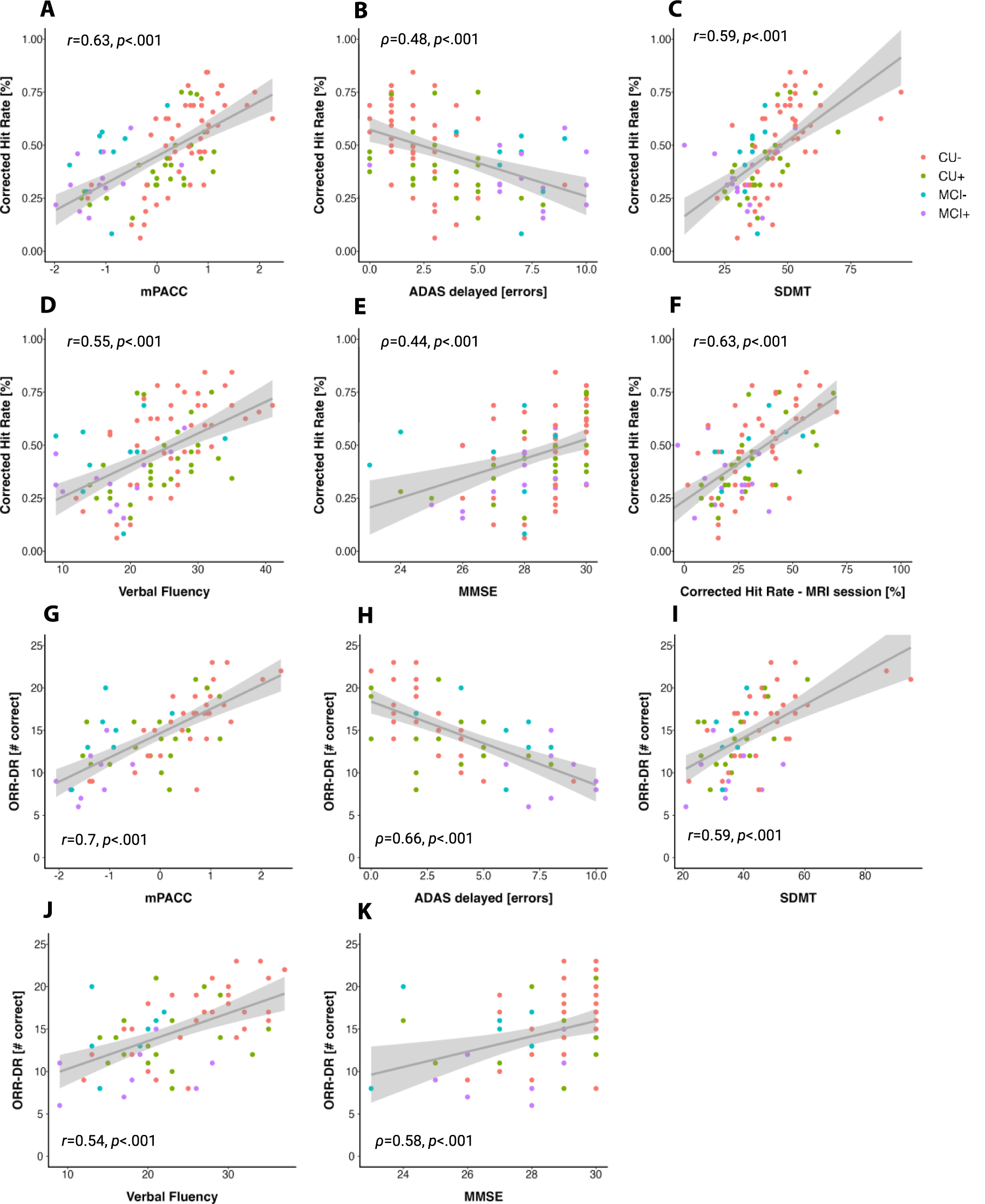
Construct validity of the Mnemonic Discrimination Task for Objects and Scenes (MDT-OS) and the Object-in-Room Recall (ORR). Scatter plots showing relationships of the MDT-OS Corrected Hit Rate with the (A) modified Preclinical Alzheimer’s Cognitive Composite, (B) errors in the delayed 10-word list recall test from ADAS-cog, (C) Symbol Digit Modalities Test, (D) Animal Fluency, (E) Mini Mental State Examination as well as the (F) Mnemonic Discrimination Task for Objects and Scenes Corrected Hit Rate from an on-site based task version that was performed during an MRI scan. Furthermore, scatter plots show relationships of the ORR Delayed Recall Score (ORR-DR) with the (G) modified Preclinical Alzheimer’s Cognitive Composite, (H) errors in the delayed 10-word list recall test from ADAS-cog, (I) Symbol Digit Modalities Test, (J) Animal Fluency and the (K) Mini Mental State Examination.

The mPACC score was associated with MDT-OS (mPACC: *r*=0.63, *p*<.001; MDT-O: *r*=0.61, *p*<.001; MDT-S: *r*=0.51, *p*<.001) and ORR delayed retrieval performance (*r*=0.7, *p*<.001). All relationships above survived corrections for multiple comparisons using FDR correction.

### Digital memory scores are associated with measures of AD pathology and medial temporal lobe atrophy

Next, we were interested whether memory scores from remote and unsupervised assessments were associated with measures of AD pathology, namely Aβ and tau pathology, as well as measures of atrophy (see Figure 5). Both memory tasks have been designed to tax medial temporal lobe-dependent memory processes (Berron et al., 2019, 2018; Maass et al., 2019). Thus, we were interested in the relationship with measures of tau pathology representing early disease stages. Following recent findings on the relationship of memory performance with measures of MTL tau pathology and atrophy (Berron et al., 2021; Flores et al., 2022; Sanchez et al., 2021), we selected tau-PET SUVr in area 35 and the anterior and posterior hippocampus as well as median thickness of area 35 and the volume of the anterior and posterior hippocampus. Regarding Aβ, we selected on SUVr in the precuneus representing early β-amyloid deposition (Palmqvist et al., 2017).

**Figure 5:**
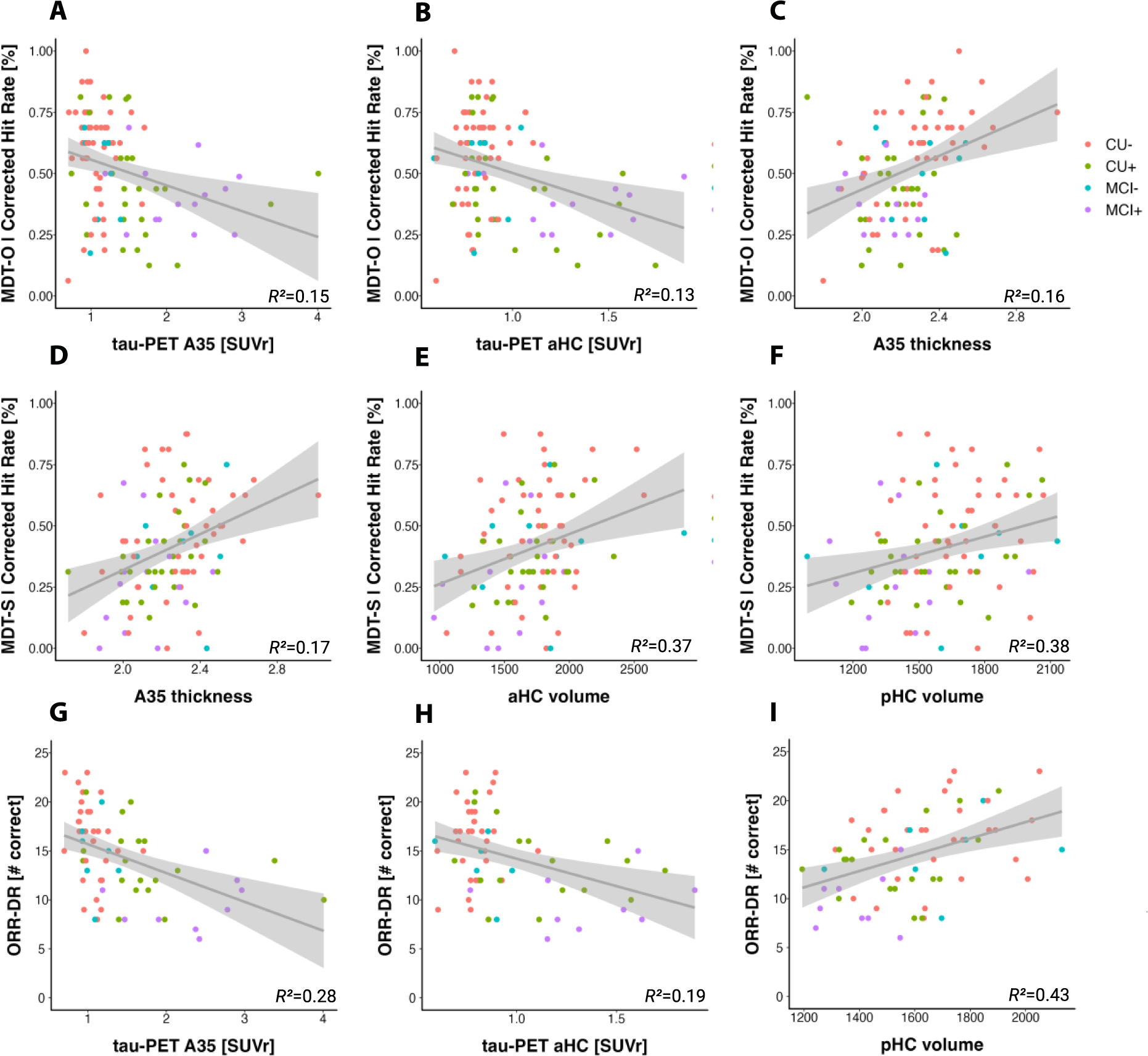
Criterion validity of the Mnemonic Discrimination Task for Objects and Scenes (MDT-OS) and the Object-in-Room Recall – Delayed Recall Score (ORR-DR). Scatter plots showing relationships between the MDT-O and tau-PET uptake in (A) area 35 and (B) the anterior hippocampus, as well as (C) cortical thickness in area 35. Panel (D), (E), and (F) show relationships of the MDT-S with area 35 cortical thickness, anterior and posterior hippocampal volume. Finally, panel (G), (H) and (I) show illustrate relationships of the ORR-DR and tau-PET uptake in area 35 and the anterior hippocampus as well as the volume of the posterior hippocampus. Abbreviations: A35, *area 35*; aHC, *anterior hippocampus*; pHC, *posterior hippocampus*; MDT-OS; *Mnemonic Discrimination task for Objects and Scenes*; ORR-DR, *Object-In-Room Recall – Delayed Recall*.

Regarding tau-PET, MDT-O, but not MDT-S, was significantly associated with A35 tau-PET SUVr (MDT-O: β=-0.71, SE=0.32, *p*=0.029; MDT-S: β=-0.32, SE=0.29, *p*=.269) and anterior hippocampal tau-PET SUVr when accounting for participant age and sex (MDT-O: β=-0.44, SE=0.14, *p*=.003; MDT-S: β=- 0.017, SE=0.13, *p*=.90). Neither measure was associated with posterior hippocampal tau-PET SUVr (MDT-O: β=-0.25, SE=0.13, *p*=.055; MDT-S: β=-0.001, SE=0.12, *p*=.966). Likewise, ORR-DR performance was significantly associated with A35 tau-PET SUVr (β=-0.06, SE=0.02, *p*=.007) and anterior (β=-0.02, SE=0.009, *p*=.016) but not posterior hippocampal tau-PET SUVr (β=-0.01, SE=0.008, *p*=.173) when accounting for participant age, sex and time between encoding and retrieval. These relationships survived corrections for multiple comparisons using FDR correction. Similarly, regarding Aβ-PET, MDT-O and ORR-DR performance, but not MDT-S, were significantly associated with Aβ-PET SUVr in precuneus (MDT-O: β=-0.29, SE=0.13, *p*=.03; MDT-S: β=-0.12, SE=0.12, *p*=.318; ORR: β=-0.02, SE=0.008, *p*=.014). However, Aβ-PET SUVr relationships did not hold when accounting for A35 tau-PET SUVR.

With respect to MTL measures of atrophy, MDT-O and MDT-S were significantly associated with area 35 thickness (MDT-O: β=0.23, SE=0.11, *p*=.043 [robust regression: F=5.34, *p*=.022]; MDT-S: β=0.22, SE=0.1, *p*=.025) but only MDT-S was associated with anterior (MDT-O: β=282.4, SE=155.4, *p*=.073; MDT-S: β=401.9, SE=135.4, *p*=.004) and posterior hippocampal volume (MDT-O: β=215.53, SE=111.6, *p*=.057; MDT-S: β=202.7, SE=99.8, *p*=.045) when accounting for age, sex and intracranial volume. ORR-DR performance was significantly associated with posterior hippocampal volume (β=14.8, SE=5.7, *p*=.013) but neither anterior hippocampal volume (β=14.7, SE=7.8, *p*=.064) nor area 35 thickness (β=0.008, SE=0.006, *p*=.175) when accounting for age, sex, intracranial volume and time between encoding and retrieval. The relationships of the MDT-O and MTL atrophy measures did not survive corrections for multiple comparisons using FDR correction.

Thus, both memory tasks showed sensitivity to measures of AD pathology as well as MTL atrophy as a measure of neurodegeneration.

### Combination of blood-based biomarker and digital cognitive marker predicts future cognitive decline

Next, we examined whether baseline scores from unsupervised memory assessments were associated with future cognitive decline rates in the mPACC and whether a combination of a blood-based biomarker and digital cognitive markers would outperform individual measures in predicting future cognitive decline. Recent work has shown that plasma p-tau217 was associated with cognitive decline as well as disease progression (Mattsson-Carlgren et al., 2023; Palmqvist et al., 2021). To that end, we derived participant-specific mPACC slopes from linear mixed-effects model with random intercepts and slopes and ran hierarchical linear regression models with plasma p-tau217 and remote memory measures as predictors in order to test which model best predicted future mPACC decline across 5 years.

Adjusting for covariates, cognitive outcomes for MDT-S and ORR-DR as well as p-tau217 but not MDT-O were significantly associated with mPACC slopes (see Table 2 and 3, see also supplementary table 1 for individual longitudinal linear mixed-effects models for each marker).

**Table 2:** Results of multiple regression models in the entire sample (n=85) predicting future cognitive decline including baseline plasma and digital remote markers as predictors, demographic covariates, and mPACC slopes as an outcome.

|  | Null model |  |  | Digital Cognitive Marker |  |  | BBM |  |  | Digital Cognitive Marker + BBM |  |  |
| --- | --- | --- | --- | --- | --- | --- | --- | --- | --- | --- | --- | --- |
| Predictor | Estimate | CI | p | Estimate | CI | p | Estimate | CI | p | Estimate | CI | p |
| (intercept) | 0.250 | 0.170 – 0.330 | <0.001 | 0.092 | -0.003 – 0.188 | 0.058 | 0.244 | 0.168 – 0.319 | <0.001 | 0.126 | 0.045 – 0.206 | 0.002 |
| Age [years] | -0.004 | -0.005 – -0.003 | <0.001 | -0.002 | -0.003 – -0.001 | <0.001 | -0.003 | -0.004 – -0.002 | <0.001 | -0.002 | -0.003 – -0.001 | <0.001 |
| Sex [female] | 0.022 | 0.003 – 0.041 | 0.020 | 0.041 | 0.022 – 0.060 | <0.001 | 0.024 | 0.006 – 0.042 | 0.008 | 0.039 | 0.021 – 0.057 | <0.001 |
| Education [years] | 0.002 | -0.001 – 0.005 | 0.256 | -0.001 | -0.004 – 0.002 | 0.574 | 0.001 | -0.002 – 0.004 | 0.566 | -0.001 | -0.004 – 0.001 | 0.307 |
| MDT-O |  |  |  | 0.040 | -0.019 – 0.099 | 0.179 |  |  |  |  |  |  |
| MDT-S |  |  |  | 0.152 | 0.098 – 0.206 | <0.001 |  |  |  | 0.157 | 0.111 – 0.203 | <0.001 |
| p-tau217 |  |  |  |  |  |  | -0.028 | -0.035 – -0.021 | <0.001 | -0.026 | -0.034 – -0.019 | <0.001 |
| Observations | 537 |  |  | 537 |  |  | 537 |  |  | 537 |  |  |
| R<sup>2</sup>/R<sup>2</sup> adj | 0.144 / 0.139 |  |  | 0.217 / 0.210 |  |  | 0.226 / 0.221 |  |  | 0.287 / 0.281 |  |  |
| AIC | -846.013 |  |  | -890.164 |  |  | -898.508 |  |  | -940.504 |  |  |

We next aimed to define an optimal combination to predict mPACC slopes. The best combination model for prediction of mPACC slopes (i.e., the model with the lowest AIC) included plasma p-tau217 (β [SE] = −0.026 [0.004]; *P* < .001), MDT-S (β [SE] = 0.157 [0.023]; *P* < .001), age (with higher age associated with worse mPACC slopes; β [SE] = -0.002 [0.001]; *P* = < .001), sex (with male sex associated with worse mPACC slopes; β [SE] = 0.039 [0.009]; *P* < .001), and education (β [SE] = -0.001 [0.001]; *P*= .307; AIC for the overall model: -941; *R*^2^ = 0.29, see Table 2 “Digital Cognitive Marker + BBM”). This model was also better than both the Digital Cognitive Marker and the BBM model as indicated by a lower AIC.

Similarly, in a smaller sample of individuals that completed both the MDT-OS and the ORR-DR (see supplementary table 2 for sample characteristics), the best combination model for prediction of mPACC slopes included plasma p-tau217 (β [SE] = −0.03 [0.004]; *P* < .001), ORR-DR (β [SE] = 0.012 [0.002]; *P* < .001), MDT-S (β [SE] = 0.142 [0.029]; *P* < .001), age (β [SE] = 0.001 [0.001]; *P* = .252), sex (with male sex associated with worse mPACC slopes; β [SE] = 0.031 [0.012]; *P* < .001), and education (with higher education associated with worse mPACC slopes; (β [SE] = -0.006 [0.002]; *P* < .001; AIC for the overall model: -606; *R*^2^ = 0.38, see Table 3 “Digital Cognitive Marker + BBM”). This model was also better than both the Digital Cognitive Marker and the BBM model with respect to lower AIC.

**Table 3:**
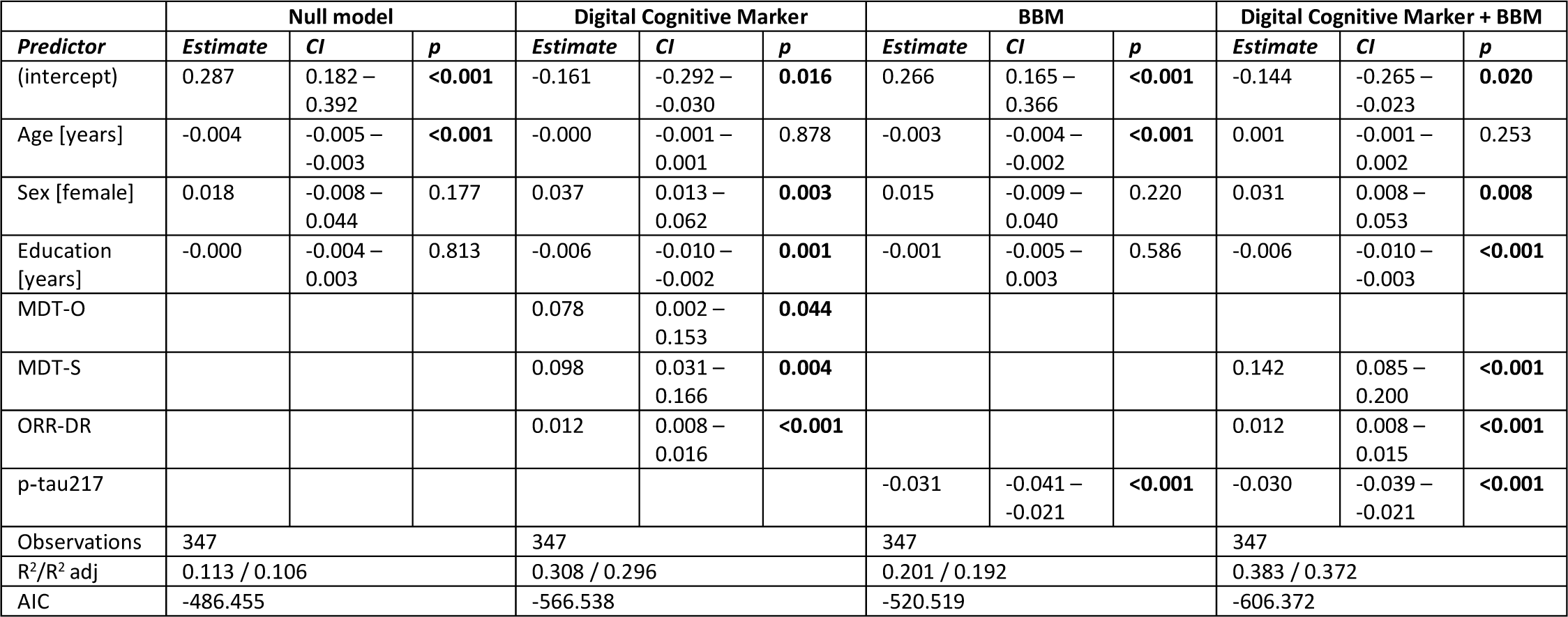
Results of multiple regression models in a restricted sample (n=66) where both digital markers were completed predicting future cognitive decline including baseline plasma and digital remote markers as predictors, demographic covariates, and mPACC slopes as an outcome.

### Remote and unsupervised short assessments show moderate-to-good retest reliability

Finally, we were interested in retest reliability of remote and unsupervised assessments given that unsupervised and remote memory assessments are well suited to assess longitudinal memory trajectories. To that end we limited the dataset to individuals that had at least four sessions completed and calculated two-way random intraclass correlation coefficients (ICC) for two scenarios. Regarding a scenario where single test sessions would be used, we calculated the ICC between test session 1 and 2 and regarding a scenario where the average between two sessions would be used, we calculated the ICC between the average of test session 1 and 2 and the average of test session 3 and 4. A single session of MDT-OS (ICC of 0.64, 95% CI [0.48, 0.76]; MDT-O: 0.57 [CI 0.39, 0.71]; MDT-S: 0.49 [CI 0.29, 0.65]; n=71) and a single session of ORR (ICC of 0.64 [CI 0.4, 0.8]; *n*=38) both showed moderate retest reliability. In the second scenario, the average of two test sessions of MDT-OS (ICC of 0.79 [CI 0.68, 0.87]; MDT-O: 0.64 [CI 0.48, 0.76]; MDT-S: 0.62 [CI 0.41, 0.76]), as well as the average of two test sessions of ORR (ICC of 0.81 [CI 0.67, 0.9]) showed good retest reliability.

## Discussion

We found construct validity of remote and unsupervised digital memory assessments to be good when comparing them with in-clinic traditional neuropsychological assessments, and that baseline performance in the remote memory assessments was associated with future decline in the mPACC score. The model that best predicted future cognitive decline in the mPACC included a combination of plasma p-tau217 and MDT-S. In a restricted sample of individuals where both digital markers were completed, the model that best predicted future cognitive decline in the mPACC also included the ORR-DR. Retest reliability was moderate-to-good when repeating the tests utilizing parallel test versions. In addition, digital memory assessments were significantly associated with PET and CSF measures of tau pathology as well as downstream MRI measures of MTL atrophy. Finally, the onboarding to the study as well as the unsupervised completion of memory tasks itself was rated positively by participants via questionnaires.

### Older adults and patients were able to complete unsupervised digital assessments

There exist stereotypes regarding older adults’ and patients’ familiarity with smartphones and tablet computers and their willingness to participate in remote and unsupervised studies using mobile devices. However, recent work has shown that, while older adults were indeed less familiar with technology, the majority of older participants decided to participate in remote studies and showed exceptional adherence when studies were planned thoughtful and included user-centered design (Nicosia et al., 2022a). In line with this, we found that a large majority of elderly participants owned a smartphone or tablet and downloaded apps by themselves or with help. The participants rated both of the current remote memory assessments as challenging but not too difficult. However, while it was intended that participants perform the ORR with a delay period of 60 minutes, there was high variability of delay times due to participants postponing the recall tasks for too long. Given that the delay period is a critical factor for task difficulty, we thus excluded all sessions with delay periods longer than 4 hours which resulted in a mean delay period of 96 minutes. Future studies should incorporate appropriate counter measures to minimize this delay period.

The main reason to not participate in the smartphone study was the high frequency of test sessions. However, the sub-sample of participants who finished the study after one year found that the number of tests was just right. A large majority found the instructions clear and the application easy to use. Three-quarters even indicated they would prefer remote mobile cognitive tests over traditional in person paper-pencil assessments. Overall, the app was rated seven out of ten by more than 80% of participants. However, participants that dropped out of the study rated the app lower and found that the number of tests were too high and durations were too long. Overall this indicates that the app was usable and easy to work with for our study participants, but that the intense continuous study schedule might be optimized.

### Remote and unsupervised assessments reflect traditional neuropsychological assessments

When introducing a novel cognitive measure, its construct validity needs to be assessed by comparing it against established neuropsychological measures of constructs it is intended to measure. This is particularly true for unsupervised and remote digital assessments that are completed in an unstandardized environment of the participant’s choice. Recent work using smartphone-based cognitive assessments in samples of older participants could successfully demonstrate high construct validity across various cognitive domains (Nicosia et al., 2022b; Öhman et al., 2022; Papp et al., 2021; Singh et al., 2023; Vyshedskiy et al., 2022). Both the ORR and the MDT-OS are intended to measure memory. While the ORR specifically aims to measure delayed memory, the MDT-OS aims to measure precision memory in an n-back task design known to also rely on attentional and executive processes (Gajewski et al., 2018). Accordingly, we found that while the ORR-DR was most strongly associated with ADAS delayed recall and less with SDMT, verbal fluency and the MMSE, the MDT-OS was associated with SDMT, ADAS delayed recall and verbal fluency and less so with the MMSE. The strongest relationship for both measures, however, was found with the mPACC. These findings nicely support earlier work using the neotiv memory tasks in samples with cognitively unimpaired individuals and MCI patients (Berron et al., 2023; Öhman et al., 2022). Furthermore, baseline performance in the MDT-S and the ORR-DR was associated with future decline in the mPACC in this study, demonstrating first evidence of its prognostic validity regarding future cognitive decline.

While high construct validity indicates that the ORR and the MDT-OS were successfully designed to measure memory function and in part executive and attentional processes, it also indicates that study participants were seemingly successful in completing the cognitive tests in an appropriate environment. While we do not know details about the individual test environment, our findings indicate that it was generally an undisturbed environment as 88% of all test sessions were reported to be without any distractions. Interestingly, our findings show that cognitively unimpaired individuals reported more distractions compared to MCI patients, in line with earlier research (Madero et al., 2021).

### Remote and unsupervised assessments are sensitive to measures of AD pathology

Both the MDT-OS and the ORR are considered MTL-dependent tasks (Berron et al., 2019, 2018; Grande et al., 2022, 2021; Maass et al., 2019) and earlier work has already linked performance in the MDT-OS with fluid and imaging measures of AD pathology (Berron et al., 2019; Maass et al., 2019). In line with these earlier findings, we found that MDT-O and ORR-DR were both significantly associated with tau-PET measures from the hippocampus and area 35, two brain regions that are affected by tau pathology very early on in the course of AD (Berron et al., 2021). Also, in line with earlier work, we did not observe such a relationship for the MDT-S (Berron et al., 2019). Finally, we found that remote digital assessments were associated with MTL atrophy, which is considered a downstream effect of AD pathology. Area 35 and the hippocampus are among the earliest sites of atrophy in AD (Berron et al., 2021; Xie et al., 2020, 2018). While the MDT-OS was associated with atrophy of the hippocampus and area 35, ORR-DR was only associated with posterior hippocampal atrophy. Taken together, this shows that both tasks depend on the functional integrity of the MTL and task performance is affected by early accumulation of AD pathology.

### Potential of remote assessments for case finding, monitoring and prognosis of cognitive impairment **in Alzheimer’s disease**

Remote and unsupervised digital cognitive assessments hold promise for case finding in health care and clinical trials, but also for longitudinal cognitive monitoring and even prognosis of cognitive decline and disease progression. Recent work showed that outcomes from remote assessments can help to identify MCI patients (Berron et al., 2023) and potentially even Aβ-positive but cognitively unimpaired participants (Jutten et al., 2022; Samaroo et al., 2020). Regarding prognosis, recent work has shown that plasma biomarkers for tau pathology are associated with future cognitive decline (Mattsson-Carlgren et al., 2023), and in combination with brief in-clinic cognitive tests, can identify MCI patients who are likely to progress towards dementia (Palmqvist et al., 2021). High construct validity and first evidence of prognostic validity regarding cognitive decline in the mPACC suggest that remote memory assessments may also have potential utility in prognosis. Indeed, our analysis comparing models with plasma p-tau217 against a model combining plasma p-tau217 and remote digital memory assessments suggests that incorporating remote digital memory assessments can significantly contribute to the prediction of future decline. Moreover, the moderate-to-good retest reliability of both remote memory assessments, coupled with the minimal practice effects due to the utilization of parallel test sets (Berron et al., 2022) hints that these tests might prove beneficial in monitoring cognitive change over time. While retest-reliability was moderate when using one single assessment, it was good when averaging across only two sessions. Future studies should investigate whether longitudinal trajectories derived from remote and unsupervised cognitive assessments can capture subtle cognitive decline.

We need to carefully consider some limitations to this study. Firstly, the sample size is relatively modest, particularly for participants who completed both digital remote and unsupervised memory paradigms. Secondly, our implementation of the ORR did not strictly enforce adherence to the planned retrieval delay intervals, which led some individuals to perform recall assessments after prolonged delays. Given the impact of delay length on task performance, we excluded sessions with significantly extended delay periods (more than 240 minutes) resulting in substantial reduction of test sessions in this study. Thus, future implementations of this task, and remote and unsupervised assessments of long-term memory in general, should make it easier for participants and patients to integrate remote and repeated tests into their everyday life, while still enforcing minimized delay periods.

## Conclusion

Our results demonstrate that unsupervised and remote digital memory assessments could effectively become a valuable tool in the diagnosis and prognosis of AD, conceivably in combination with plasma biomarkers.

## Data Availability

Anonymized data will be shared by request from a qualified academic investigator for the sole purpose of replicating procedures and results presented in the article and as long as data transfer is in agreement with EU legislation on the general data protection regulation and decisions by the Ethical Review Board of Sweden and Region Skane, which should be regulated in a material transfer agreement.

## Acknowledgements

We want to thank all participants of the Swedish BioFINDER-2 study and their families for their participation in the study. We also want to thank Oisín Clancy and Hannah Baumeister for their help with the manual quality assessment of the medial temporal subregional masks.

