## Supplementary material for "Remote and unsupervised digital memory assessments can reliably detect cognitive impairment in Alzheimer’s disease"

**Supplementary table 1: Longitudinal random slope linear mixed models in the entire sample predicting future cognitive decline including baseline plasma and digital remote markers and years since baseline as predictors, demographic covariates, and mPACC slopes as an outcome. The term of interest is the interaction between baseline markers and years since baseline.**

| | **MDT-S** | | | **MDT-O** | | | **ORR-DR** | | | **p-tau217** | | |
|---|---|---|---|---|---|---|---|---|---|---|---|---|
| ***Predictor*** | ***Estimate*** | ***CI*** | ***p*** | ***Estimate*** | ***CI*** | ***p*** | ***Estimate*** | ***CI*** | ***p*** | ***Estimate*** | ***CI*** | ***p*** |
| (Intercept) | 0.658 | -0.689 – 2.006 | 0.337 | -0.373 | -1.724 – 0.977 | 0.587 | -1.224 | -3.147 – 0.699 | 0.211 | 1.757 | 0.370 – 3.144 | **0.013** |
| YearsSince Baseline | -0.166 | -0.275 – -0.058 | **0.003** | -0.149 | -0.293 – -0.004 | **0.043** | -0.302 | -0.532 – -0.072 | **0.010** | 0.042 | -0.021 – 0.105 | 0.195 |
| MDT-S | 1.440 | 0.707 – 2.174 | **<0.001** | | | | | | | | | |
| diagnosis [CU+] | -0.327 | -0.675 – 0.021 | 0.066 | -0.159 | -0.487 – 0.169 | 0.340 | -0.401 | -0.821 – 0.020 | 0.062 | -0.071 | -0.480 – 0.339 | 0.735 |
| diagnosis [MCI-] | -1.480 | -1.980 – -0.981 | **<0.001** | -1.395 | -1.865 – -0.926 | **<0.001** | -1.446 | -1.997 – -0.895 | **<0.001** | -1.565 | -2.122 – -1.007 | **<0.001** |
| diagnosis [MCI+] | -1.665 | -2.099 – -1.232 | **<0.001** | -1.445 | -1.856 – -1.034 | **<0.001** | -1.348 | -1.925 – -0.772 | **<0.001** | -1.237 | -1.903 – -0.571 | **<0.001** |
| age | -0.021 | -0.037 – -0.005 | **0.010** | -0.016 | -0.031 – -0.001 | **0.034** | -0.012 | -0.032 – 0.007 | 0.218 | -0.033 | -0.049 – -0.016 | **<0.001** |
| sex [female] | 0.128 | -0.157 – 0.413 | 0.379 | 0.213 | -0.057 – 0.483 | 0.122 | 0.053 | -0.295 – 0.401 | 0.763 | 0.062 | -0.260 – 0.384 | 0.705 |
| Education [years] | 0.020 | -0.024 – 0.064 | 0.374 | 0.022 | -0.019 – 0.063 | 0.283 | 0.026 | -0.025 – 0.077 | 0.309 | 0.049 | -0.003 – 0.100 | 0.062 |
| YearsSince Baseline * MDT-S | 0.245 | 0.010 – 0.481 | **0.041** | | | | | | | | | |
| MDT-O | | | | 2.211 | 1.447 – 2.976 | **<0.001** | | | | | | |
| YearsSince Baseline * MDT-O | | | | 0.152 | -0.109 – 0.413 | 0.253 | | | | | | |
| ORR-DR | | | | | | | 0.121 | 0.069 – 0.173 | **<0.001** | | | |
| YearsSince Baseline * ORR-DR | | | | | | | 0.017 | 0.002 – 0.032 | **0.027** | | | |
| p-tau217 | | | | | | | | | | -0.101 | -0.252 – 0.049 | 0.187 |
| YearsSince Baseline * p-tau217 | | | | | | | | | | -0.042 | -0.076 – -0.009 | **0.013** |
| Random Effects | | | | | | | | | | | | |
| $\sigma^2$ | 0.196 | | | 0.192 | | | 0.176 | | | 0.157 | | |
| $\tau_{00}$ | 0.344 UserID | | | 0.271 UserID | | | 0.301 UserID | | | 0.421 UserID | | |
| $\tau_{11}$ | 0.035 UserID.YearsSinceBaseline | | | 0.040 UserID.YearsSinceBaseline | | | 0.035 UserID.YearsSinceBaseline | | | 0.014 UserID.YearsSinceBaseline | | |
| $\rho_{01}$ | 0.094 UserID | | | 0.231 UserID | | | 0.133 UserID | | | 0.145 UserID | | |
| ICC | 0.751 | | | 0.756 | | | 0.758 | | | 0.780 | | |
| N | 99 UserID | | | 99 UserID | | | 65 UserID | | | 85 UserID | | |
| Observations | 325 | | | 325 | | | 211 | | | 276 | | |
| Marginal $R^2$ / Conditional $R^2$ | 0.545 / 0.887 | | | 0.552 / 0.891 | | | 0.623 / 0.909 | | | 0.488 / 0.887 | | |
| AIC | 717.263 | | | 707.624 | | | 472.243 | | | 565.454 | | |

**Supplementary table 2: Demographics of the sample that performed the MDT-OS and ORR-DR**

| | **CU Aβ- (N=31)** | **CU Aβ+ (N=19)** | **MCI Aβ- (N=7)** | **MCI Aβ+ (N=9)** | **Total (N=66)** |
|---|---|---|---|---|---|
| *Age (years)* | 61.3 (10.9) | 71.4 (7.4) | 64 (13.1) | 67.8 (4.3) | 65.4 (10.4) |
| *Education (years)* | 13.1 (3.1) | 12.8 (3.6) | 12.7 (3) | 13.6 (4) | 13 (3.3) |
| *Sex (% female)* | 51.6 % | 68.4 % | 71.4 % | 22.2 % | 54.5 % |
| *MMSE* | 29 (1.2) | 28.3 (1.8) | 26.4 (2.1) | 27.6 (1.5) | 28.3 (1.7) |
